# Evaluating treatment strategies for managing anaemia with erythropoiesis stimulating agent therapy in haemodialysis patients: findings from a target trial emulation using electronic health record data

**DOI:** 10.1101/2025.10.17.25338210

**Authors:** Kate Birnie, Fergus J Caskey, Yoav Ben-Shlomo, Dorothea Nitsch, Anna Casula, Eleanor J Murray, Jonathan AC Sterne

## Abstract

**Background:** Anaemia is a common complication in chronic kidney disease (CKD), often managed with erythropoiesis-stimulating agents (ESAs) and intravenous iron. However, the optimal haemoglobin (Hb) target for treatment remains uncertain. We used electronic health records to emulate a target trial comparing different Hb targets in haemodialysis patients, including ranges used in practice and those tested in earlier trials, to estimate their effects on mortality.

**Methods:** We used electronic health records from selected UK kidney centres reporting to the Renal Registry. Eligible patients were ≥18 years, on haemodialysis for ≥3 months, and without marked ESA resistance. Using the clone-censor-weight method, we emulated a trial comparing Hb targets of (1) 105–125 g/L versus 95–115 g/L, and (2) 125–145 g/L versus 95–115 g/L. Darbepoetin dosing followed a standardized protocol with predefined dose adjustment rules and a maximum of 150 µg/week. The outcome was all-cause mortality over 8 months.

**Results:** Among 8,628 patients from 10 kidney centres followed between 2004 and 2016; 62% were male, and the median age was 66 years (IQR 52–76). At baseline, mean Hb was 97.7 g/L (SD 15.1) and 79% were receiving darbepoetin (median dose 30 µg/week). The estimated hazard ratios for mortality were 0.92 (95% CI 0.75, 1.12) comparing the 105–125 g/L and 95–115 g/L Hb target ranges, and HR 1.20 (95% CI 0.94, 1.54) comparing the 125–145 g/L and 95–115 g/L ranges. Fewer patients remained adherent to the 125–145 g/L target range, limiting precision.

**Conclusions:** Compared with targeting a Hb range of 95-115 g/L Hb, targeting a slightly higher range of 105– 125 g/L under controlled ESA dosing was not associated with increased mortality in haemodialysis patients. However, targeting Hb levels above 125 g/L may increase mortality risk, consistent with previous trial findings.

**Lay Summary:** Anaemia, or low blood haemoglobin levels, is common in people receiving dialysis for kidney disease. It is usually treated with medicines that help the body make red blood cells, along with iron therapy. But the optimal haemoglobin target is unknown. Past clinical trials did not show clear benefits from targeting higher levels and sometimes found increased risks of heart problems, so new trials are unlikely to take place. We designed a “target trial” to mimic a randomized trial using existing health records. We used data from over 8,000 UK dialysis patients between 2004 and 2016 to emulate a trial. Aiming for a moderate haemoglobin range (105–125 g/L) did not increase the risk of death compared to a lower range (95–115 g/L), whereas aiming above 125 g/L may increase risk. These findings suggest that modestly higher haemoglobin targets, if carefully managed, could improve symptoms and quality of life without increasing harm.

## Introduction

Anaemia, defined by the World Health Organisation as haemoglobin (Hb) levels <130 g/L in men and <120 g/L in non-pregnant women^1^, leads to reduced quality of life^2^, fatigue^3^, decreased exercise capacity^4^ and shortness of breath^5^ in individuals with chronic kidney disease (CKD) and end-stage kidney disease who require dialysis.^6^ Erythropoiesis stimulating agents (ESAs), along with intravenous iron supplementation, are used to increase Hb levels, but the optimal level of Hb that should be targeted remains unknown. UK clinical guidelines recommend a target Hb of 100-120 g/L^7^ and Kidney Disease Outcomes Quality Initiative (KDOQI) guidelines recommend Hb should generally be in a target range of range of 110 to 120 g/L and the target should not be greater than 130 g/L.^8^ A 2025 update to the Kidney Disease: Improving Global Outcomes (KDIGO) guideline (currently in draft and under public review) recommends targeting Hb levels below 115 g/L.^9^

Randomized trials in patients with CKD^10–12^ and end-stage kidney disease^13^ found either no change or an increase in the risk of cardiovascular events with higher Hb targets. However, reaching a target Hb of 120 g/L requires high doses of ESA in some individuals and high ESA doses are associated with a higher risk of death in observational studies and post hoc analyses of randomized trials.^14–16^ It has been proposed that high ESA doses can cause thrombosis^17^ in the presence of relative iron deficiency, as well as arterial hypertension, endothelial activation, increased platelet reactivity, increased blood coagulability, and accelerated tumour growth, and inflammation.^18–21^ As a result, commonly used Hb targets have an upper limit of 120 g/L.

It is also possible that the higher risks associated with high ESA doses simply reflect that ESA resistance is a marker of other co-morbidities (inflammation and cardiovascular disease). If so, individuals without ESA resistance may benefit from a higher Hb target than the one typically used. Because it is unlikely that new randomized trials will be conducted, this question needs to be addressed using observational data. We used UK electronic health record data to emulate a target trial comparing the effects of ESA treatment strategies on mortality in haemodialysis patients. The target ranges compared were (1) 105–125 g/L versus 95–115 g/L and (2) 125–145 g/L versus 95–115 g/L.

## Methods

### Data

The UK Renal Registry (UKRR) collects clinical and biochemical data for all UK patients aged ≥18 years receiving kidney replacement therapy (KRT).^22^ A bespoke data extraction was obtained for selected kidney centres reporting to the UKRR. In each quarter of a calendar year, centres reporting <60% of haemodialysis patients being treated with erythropoiesis-stimulating agents (ESAs) were considered to have incomplete data so were excluded. Data were extracted from 2004 to 2016, though the contribution period varied by centre based on when prescribing data were sufficiently complete. Analyses were restricted to patients prescribed darbepoetin, as dose equivalency with other ESAs (e.g., epoetin) was insufficiently similar to allow combined analyses.

### Research Ethics and Informed Consent

The processing of UKRR data for research has been approved by the NRES Committee North East Newcastle and North Tyneside 1 Research Ethics Committee, reference 21/NE/0045. The UKRR has section 251 permissions to use data for research without individual patient consent.

### Emulating a target trial

The target trial emulation and analysis methodology has been described in detail previously.^23^ Patients were eligible if they were ≥18 years old and had been on haemodialysis for ≥3 months. We excluded patients with evidence of marked ESA resistance (operationalised as receiving a high darbepoetin dose [≥120 µg/week]) and with low Hb [<80 g/L]) at baseline. The outcome was all-cause mortality over 8 months.

We first compared Hb target range 105–125 g/L with range 95–115 g/L. These ranges were chosen to fall slightly above and below the UK guideline range of 100–120 g/L^7^ while reflecting the spectrum of Hb targets recommended or observed in international guidelines. We also compared target range 125–145 g/L with range 95–115 g/L. The 125–145 g/L Hb range was chosen to align with near-complete correction of anaemia, consistent with historical randomized trials in which Hb targets approached or exceeded 130 g/L.^10–13^

Darbepoetin dose adjustments followed a standardized protocol designed to reflect typical clinical practice. Decisions to modify doses were based on current and recent Hb levels, whether the dose had been changed in the previous month, and observed Hb response to prior dosing adjustments (Figure 1). Dose changes were allowed only within a predefined acceptable range, with a maximum dose of 150 µg/week permitted (Supplementary Figure A1). Patients showing signs of ESA resistance requiring dose escalations were censored unless the dose remained within the allowed limits. Additionally, a grace period (within which changes should be made) of up to one month was incorporated to reflect delays in dose implementation. This grace period helped accommodate treatment variability and minimized censoring caused by minor deviations from the dosing rules.

**Figure 1.**
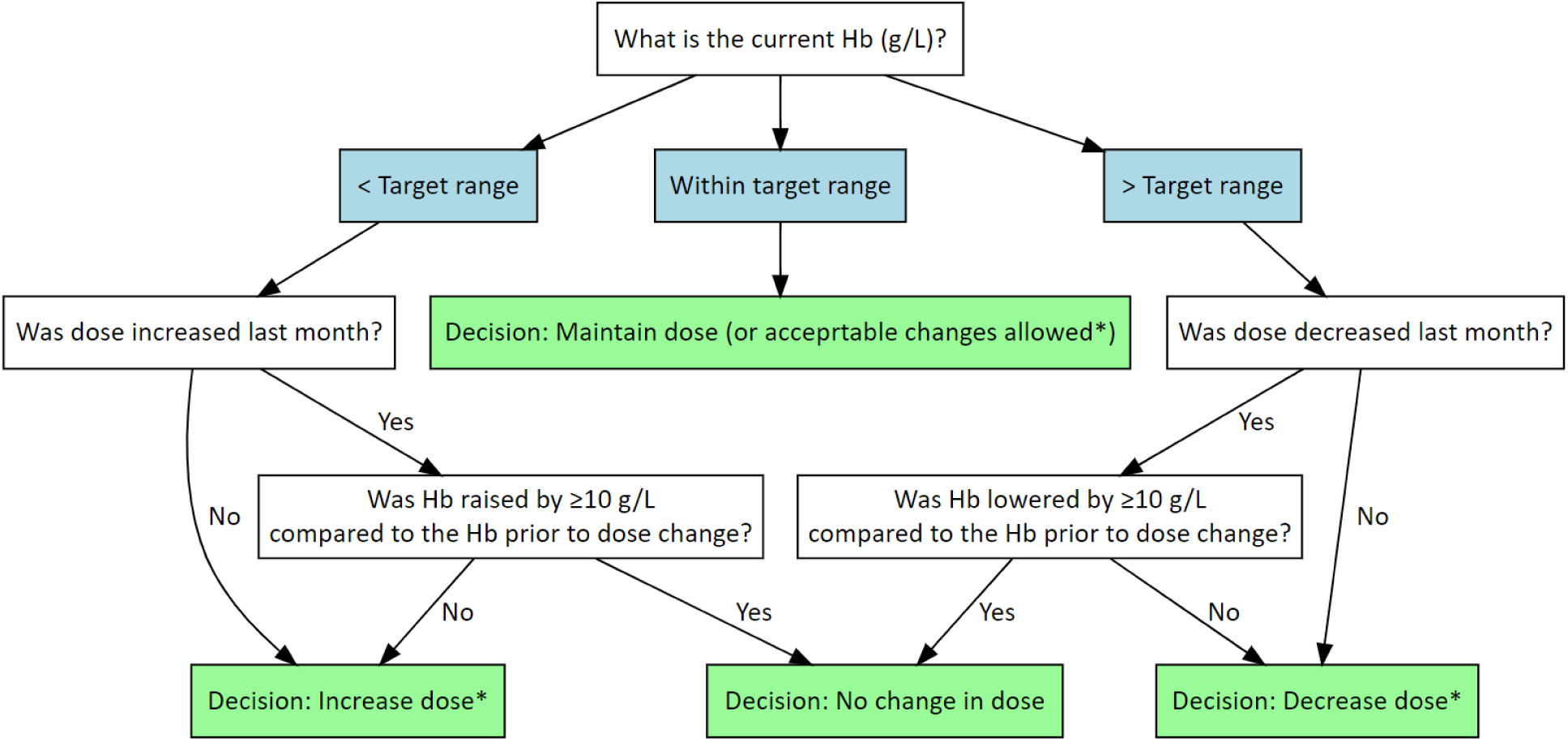
Dose decision protocol Notes: Hb: haemoglobin. * When the decision is to change the dose, the new dose needs to be within acceptable levels (see Figure A1). The figure has been adapted from Birnie et al., *Epidemiology* 2023;34:879– 887.^23^

We used the three-step ‘clone, censor, inverse-probability weight’ framework for comparing sustained treatment strategies^24^. 1. Cloning: All patients met both strategies’ eligibility criteria at baseline, defined as the latest of the dates when their dialysis centre became eligible and when they first met the eligibility criteria. At this point (time zero of the target trial), their follow-up was duplicated, with one copy (‘clone’) assigned to each strategy being compared. 2. Censoring: Follow-up ended when the patient’s data was no longer compatible with the strategy assigned to the clone because they received a darbepoetin dose outside the acceptable range for the assigned strategy. 3. Inverse probability (IP) weighting, to account for the selection bias introduced by the informative censoring in the previous step.

Data were organized into discrete 28-day intervals^25^ for each patient. Time zero was set at baseline. Each month, darbepoetin dose adjustments were recorded along with the corresponding Hb level that informed the treatment decision. Darbepoetin dose was recorded as zero if the patient was not receiving darbepoetin or as a numerical value if prescribed. Non-zero doses were log-transformed for normality. Follow-up ended eight months after baseline, death, kidney transplantation or a change to peritoneal dialysis, or loss to follow-up, whichever happened first.

Baseline covariates included: patient age (years), sex, kidney centre (the largest centre was chosen as the reference category), primary kidney disease (diabetes, glomerulonephritis, pyelonephritis, polycystic kidneys and ‘other’ [hypertension, kidney vascular disease, other, uncertain]) and co-morbidities present at the start of KRT (angina, angioplasty, claudication, chronic obstructive pulmonary disease, diabetes not causing kidney failure, ischaemic / neuropathic ulcers, liver disease, malignancy, previous myocardial infarction within last 3 months prior to starting KRT, previous myocardial infarction >3 months prior to start of KRT, previous coronary artery bypass graft or coronary angioplasty, amputation for peripheral vascular disease, symptomatic cerebrovascular disease, heart failure, and whether the patient was a smoker). Time-updated covariates included: Hb (g/L), darbepoetin dose (µg/week), white blood cell count (10^9^/L), albumin (g/L), ferritin (µg/L), adjusted calcium (mg/dL), C-reactive protein (mg/L), urea reduction ratio (dialysis adequacy %) number of blood tests in the previous 28 days, time since eligibility into the study (months).

### Statistical analysis

#### Deriving weights

Because follow-up for a clone was censored when patients received a darbepoetin dose outside the acceptable range for the strategy assigned to that clone, models for treatment (darbepoetin dose levels) were used to derive the probability of being censored, based on covariates up to and including the previous month. The treatment models (described in detail previously)^23^ included (1) logistic regression to estimate the probability of receiving zero darbepoetin, with separate models for patients receiving and not receiving darbepoetin in the previous month, to model cessation of darbepoetin and remaining off darbepoetin respectively; (2) a multivariable heteroskedastic linear regression model for log darbepoetin dose among those with non-zero dose; and (3) multinomial logistic regression for extreme doses (e.g. 2.5 or 150 µg/week) in that month. The multinomial models were for all possible categories of dose given the previous month. For example, from a dose of 2.5 µg/week, the possible categories are: zero dose, the same dose, an increase consistent with both strategies, an increase consistent with the higher of the Hb target strategies only, or an increase inconsistent with both strategies. Due to small sample sizes in some categories, only Hb and month were included as covariates in the multinomial models.

The probability of adhering to the assigned treatment strategy in each month was derived by combining probabilities from all relevant models. The cumulative probability of remaining uncensored was derived by multiplying the probabilities of remaining uncensored during each month since baseline, with inverse probability of censoring weights (IPCW) derived as the reciprocal of these probabilities. Patients could also be censored for other reasons: separate logistic regression models, based on covariates up to and including the previous month, were used to calculate censoring weights for patients who changed from haemodialysis to peritoneal dialysis, received a kidney transplant or were lost to follow-up and IPCW defined in the same way. IPCWs for each type of censoring were multiplied together to give the final consolidated weight for each strategy, patient and month.

Pooled logistic regression models, weighted using the IPCW for each strategy, was used to estimate the mortality hazard ratio (HR) for each comparison. These models included cubic splines with 2 knots for month (to allow for non-linear relationships) and used robust standard errors for clustering by patient. In sensitivity analyses the IPCW were truncated at the 99^th^, 95^th^ and 90^th^ percentiles (chosen a priori) to mitigate the impact of large weights. We estimated weighted survival curves under each strategy using inverse probability of treatment and censoring weights, truncated at the 99th percentile to improve stability.

Statistical analyses were conducted in Stata 17. Confidence intervals were obtained using a nonparametric bootstrap procedure with 1,000 replicates. Figures of flow diagrams and dose change visualisations were generated in R (version 4.5.1) using the DiagrammeR, ggplot2, dplyr, and tibble packages.

## Results

### Characteristics of the study cohort

A total of 8,628 patients from 10 kidney centres were eligible for the study (Supplementary Figure A2). The median age was 66 years (IQR 52–76) and 5,351 patients (62%) were male (Table 1). Diabetes and glomerulonephritis were the primary kidney diseases in 1,783 (20.7%) and 1,228 (14.2%) patients respectively. At baseline 6,773 (78.5%) patients were being treated with darbepoetin, with a median dose of 30 µg/week (IQR 20, 50) and a mean Hb of 97.7 g/L (SD 15.1).

**Table 1.** Characteristics of study participants at baseline.

| **Covariate** | **Category/unit** | **N (%) or summary** |
| --- | --- | --- |
| Age (years) | Median (IQR) | 66 (52-76) |
| Sex | Male | 5351 (62.0%) |
|  | Female | 3277 (38.0%) |
| Primary kidney disease | Diabetes | 1783 (20.7%) |
|  | Glomerulonephritis | 1228 (14.2%) |
|  | Polycystic kidneys | 534 (6.2%) |
|  | Pyelonephritis | 717 (8.3%) |
|  | Miscellaneous | 4366 (50.6%) |
| Year of entering study | 2004 | 777 (9.0%) |
|  | 2005 | 737 (8.5%) |
|  | 2006 | 635 (7.4%) |
|  | 2007 | 593 (6.9%) |
|  | 2008 | 595 (6.9%) |
|  | 2009 | 570 (6.6%) |
|  | 2010 | 600 (7.0%) |
|  | 2011 | 1061 (12.3%) |
|  | 2012 | 777 (9.0%) |
|  | 2013 | 767 (8.9%) |
|  | 2014 | 640 (7.4%) |
|  | 2015 | 447 (5.2%) |
|  | 2016 | 429 (5.0%) |
| On darbepoetin treatment | Yes | 6773 (78.5%) |
|  | No | 1855 (21.5%) |
| Darbepoetin dose, for those<br>treatment (µg/week) | Median (IQR) | 30 (20, 50) |
| Hb (g/L) | Mean (SD) | 97.7 (15.1) |
| Ferritin (µg/L) | <300 | 3018 (35.0%) |
|  | 300-449 | 1545 (17.9%) |
|  | 500+ | 1501 (17.4%) |
|  | Missing | 2564 (29.7%) |
| C-reactive protein (mg/L) | 0 or not tested | 4432 (51.4%) |
|  | 0.1 to 4.9 | 426 (4.9%) |
|  | 5 to 19.9 | 1844 (21.4%) |

| Covariate | Category/unit | N (%) or summary |
| --- | --- | --- |
|  | 20+ | 1926 (22.3%) |
| Blood tests in previous month (N) | 0 | 2072 (24.0%) |
|  | 1 | 3933 (45.6%) |
|  | 2 | 853 (9.9%) |
|  | 3 | 433 (5.0%) |
|  | 4+ | 1337 (15.5%) |
| Albumin (g/L) | <35 | 4066 (47.1%) |
|  | 35-39 | 2497 (28.9%) |
|  | 40+ | 1750 (20.3%) |
|  | Missing | 315 (3.7%) |
| White blood count (10<sup>9</sup>/L) | <6 | 2316 (26.8%) |
|  | 6-6.9 | 1380 (16.0%) |
|  | 7-7.9 | 1370 (15.9%) |
|  | 8-8.9 | 1113 (12.9%) |
|  | 9+ | 2449 (28.4%) |
| Adjusted calcium (mg/dL) | <2.3 | 3667 (42.5%) |
|  | 2.3-2.39 | 1741 (20.2%) |
|  | 2.4-2.49 | 1383 (16.0%) |
|  | 2.5+ | 1428 (16.6%) |
|  | Missing | 409 (4.7%) |
| Urea reduction ratio (%) | <60 | 1692 (19.6%) |
|  | 60 to 69 | 1826 (21.2%) |
|  | 70 to 74 | 994 (11.5%) |
|  | 75-79 | 633 (7.3%) |
|  | 80+ | 320 (3.7%) |
|  | Missing | 3163 (36.7%) |

### Predictors of darbepoetin treatment and dosing

Predictors of darbepoetin dosing were consistent with clinical expectations. Prior dosing history was strongly associated with current dose, and recent rises in haemoglobin were associated with dose reductions or stopping darbepoetin. Among those already receiving darbepoetin, patients with heart failure or with frequent blood tests in the prior month were more likely to have darbepoetin stopped. Among those not receiving darbepoetin, higher ferritin and CRP levels were associated with a lower likelihood of remaining on zero dose, consistent with anaemia management decisions in the context of inflammation.

### Comparison of haemoglobin target ranges: 105–125 g/L vs 95–115 g/L

For the comparison of haemoglobin target ranges 105–125 g/L versus 95–115 g/L, Hb levels started to diverge by the second month, with the 105–125 g/L target group maintaining higher Hb levels for the remainder of follow-up (Figure 2, panel A). By month 5, mean Hb in the 105–125 g/L strategy was 114.9 g/L (95% confidence interval [CI]: 114.5, 115.4 g/L), while in the 95–115 g/L strategy, it was 111.5 g/L (111.1, 112.0 g/L). By month 5, the geometric mean darbepoetin dose (for those being treated with darbepoetin) was 39.3 µg/week (95% CI 38.4, 40.2 µg/week) in the 105–125 g/L strategy and 34.9 µg/week (95% CI 34.1, 35.7) in the 95–115 g/L strategy (Figure 2, panel C). As expected, the number of patients in the analysis reduced over time as patients stopped adhering to the assigned strategies, died, were censored or were lost to follow-up. At four months 56% patients remained in the 105–125 g/L strategy and 60% in the 95–115 g/L strategy. By eight months, 37% remained in the 105–125 g/L strategy and 35% the 95–115 g/L strategy.

**Figure 2.**
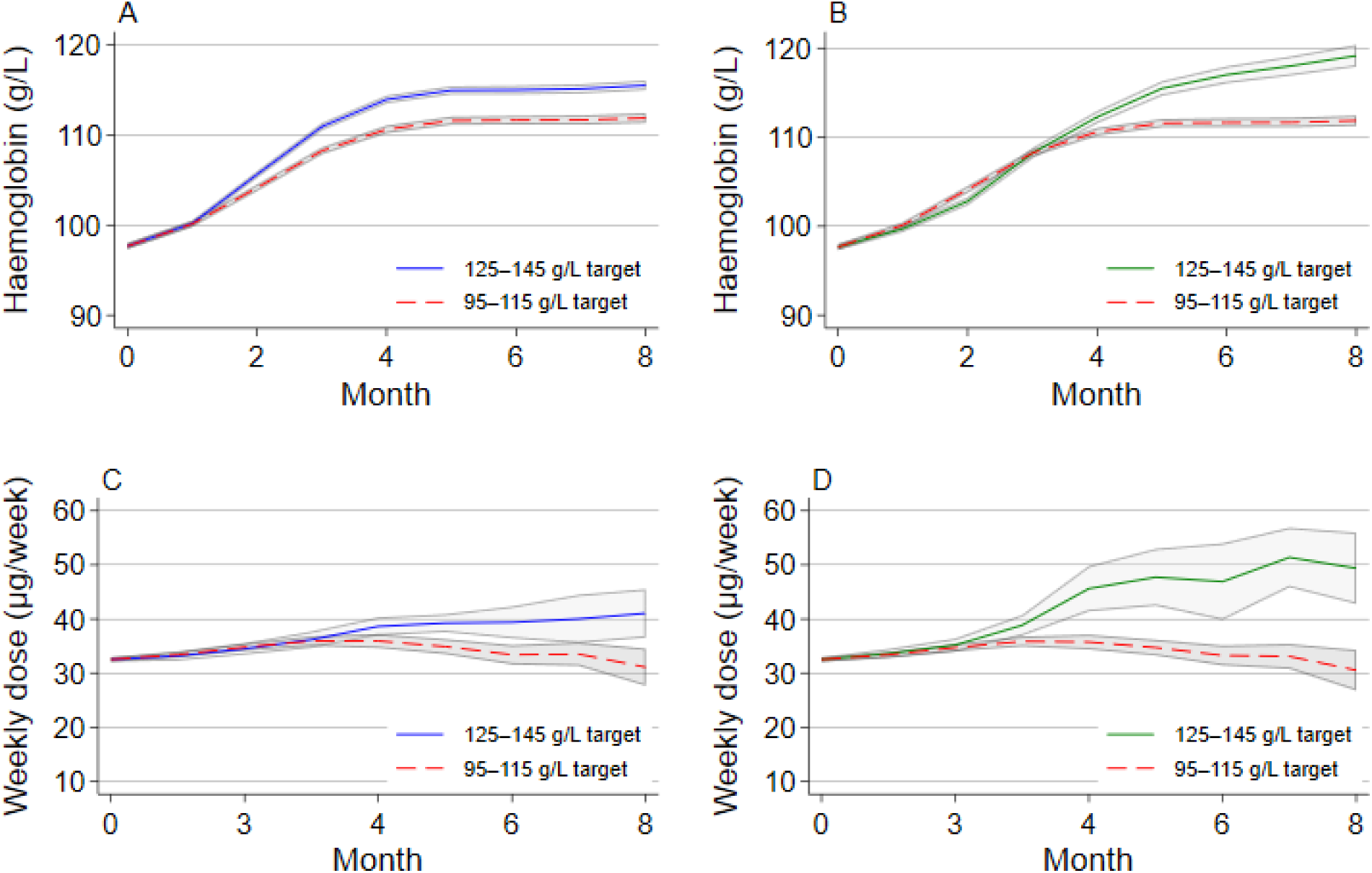
Mean Hb and geometric mean darbepoetin dose, with 95% CI, over time, by treatment strategy Geometric mean darbepoetin dose (for non-zero doses) with 95% confidence (CI) intervals from a weighted analysis

There were 373 deaths during 39,155 patient-months follow-up in the 105–125 g/L strategy group, compared to 434 deaths during 40,387 patient-months in the 95–115 g/L strategy group. In the weighted analysis, which accounted for baseline and time-updated covariates, the estimated hazard ratio (HR) comparing the 105–125 g/L versus the 95–115 g/L strategy was 0.92 (95% CI: 0.75, 1.12). The median weight was 1.5, with the 90th, 95th, and 99th percentiles at 5.2, 8.8, and 25.5, respectively. Truncating weights at the 99th, 95th, and 90th percentiles yielded similar effect estimates with narrower CIs: HRs of 0.91 (95% CI: 0.80, 1.03), 0.90 (95% CI: 0.82, 0.99), and 0.89 (95% CI: 0.82, 0.97), respectively. The unweighted estimate, which does not account for confounding, was 0.89 (95% CI 0.83, 0.94).

### Comparison of haemoglobin target ranges: 125–145 g/L vs 95–115 g/L

For the 125–145 g/L Hb strategy, mean Hb increased across follow-up, reaching 119.2 g/L (95% CI: 117.7, 120.6 g/L) by month 8 (Figure 2, panel B), when the geometric mean of darbepoetin dose (for those being treated with darbepoetin) was 49.4 µg/week (95% CI 46.6, 52.3 µg/week) (Figure 2, (Figure 2, panel D).

There were 187 deaths during 21,663 patient-months follow-up in the 125–145 g/L strategy group. Only 667 patients (8%) in the 125–145 g/L strategy group remained uncensored by month eight, indicating that this strategy was not often followed in clinical practice. The median weight was 1.5, with the 90th, 95th, and 99th percentiles at 6.2, 11.2, and 43.8, respectively.

The estimated HR comparing the 125–145 g/L with the 95–115 g/L target was 1.20 (95% CI: 0.94, 1.54) after truncating the weights at the 99^th^ percentile. Truncating weights at the 95th, and 90th percentiles progressively attenuated effect estimates and narrowed CIs: HR 1.11 (95% CI: 0.93, 1.32), and 1.03 (95% CI: 0.88, 1.21), respectively. The fully weighted model produced unstable estimates due to a small number of very large weights and is therefore not presented. The unweighted estimate was 0.95 (95% CI 0.85, 1.07).

The estimated survival curves for all three strategies are shown in Figure 3. The curve for the 125–145 g/L target strategy lies below those for the other two target ranges from around month 4 onward, although the confidence intervals are wide. Confidence intervals widened over time, reflecting greater uncertainty due to the increased proportion of patients whose follow-up was censored, reducing the amount of available data at later time points.

**Figure 3.**
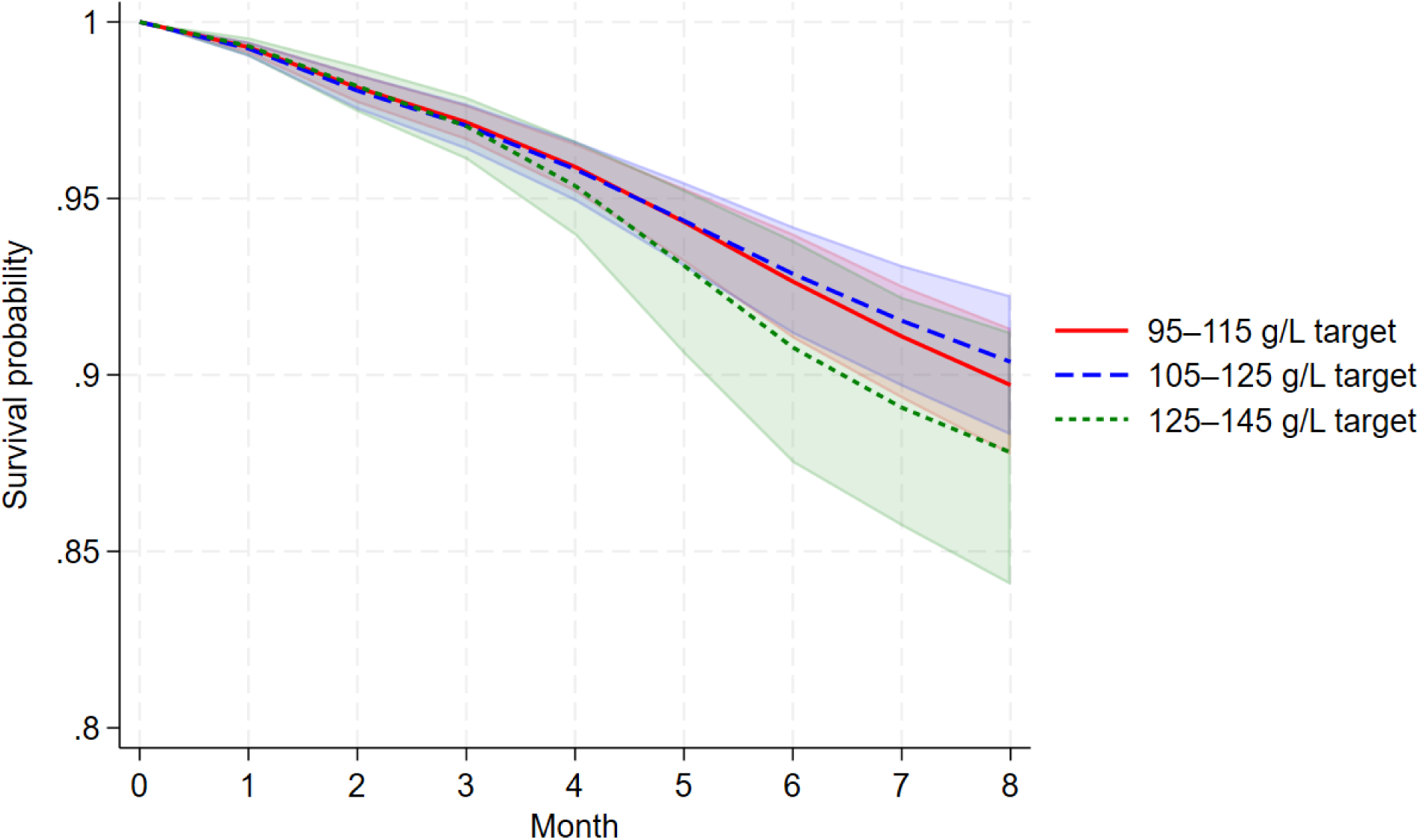
Estimated weighted survival curves for all-cause mortality for the three target Hb strategies Note: shaded areas represent 95% confidence intervals derived from bootstrap resampling

## Discussion

Using observational electronic health record data from the UKRR, we emulated target trials evaluating the effects of different Hb target strategies on mortality among haemodialysis patients. Because these strategies cannot be distinguished by patients’ characteristics at the start of follow-up, we employed the clone-censor-weight method to emulate the target trial. We found no evidence of increased all-cause mortality comparing the 105–125 g/L Hb with the 95–115 g/L target range over 8 months of follow-up. The 105–125 g/L target is slightly higher than the UK Kidney Association Clinical Practice Guidelines recommendation of 100–120 g/L^7^ and also exceeds the 115 g/L upper target limit suggested by KDIGO.^9^ The comparison of the 125–145 g/L versus 95–115 g/L target ranges suggested a potential increased mortality with the 125–145 g/L target range.

A randomized trial of 1,233 haemodialysis patients with cardiovascular comorbidity, published in 1998,^13^ was terminated early when it became clear that benefit from full correction of anaemia (compared to partial correction) was highly improbable. Randomized trials examining different Hb targets in patients with CKD include the CHOIR trial (published 2006),^12^ which randomized 1,432 patients with CKD not on KRT to Hb targets of 135 g/L or 113 g/L. Patients assigned to the higher Hb target had an increased risk (HR 1.34; 95% CI: 1.03, 1.74) of the primary composite outcome (death, myocardial infarction, hospitalization for heart failure, or stroke) compared to the lower Hb target. In contrast, the CREATE trial (published 2006),^10^ randomized 603 patients with CKD not on KRT to a target Hb of 130–150 g/L or 105–115 g/L: the HR for cardiovascular events was 0.78 (95% CI: 0.53, 1.14). The TREAT trial, published in 2009^11^, included 4,038 patients with diabetes, CKD, and anaemia. Participants were randomly assigned to achieve a haemoglobin level of approximately 130 g/L using darbepoetin alfa or placebo with rescue darbepoetin alfa if Hb <90 g/L). The HR for the composite outcome of death or cardiovascular events was 1.05 (95% CI: 0.94, 1.17), but stroke incidence was higher in the darbepoetin alfa arm (HR 1.92; 95% CI: 1.38, 2.68).

In published trials, patients assigned to higher Hb targets typically received median ESA doses two to three times greater than those assigned to lower targets.^21^ Furthermore, a secondary analysis of CHOIR study data found that failure to reach the target Hb and high ESA doses were associated with a higher risk of primary outcomes, including death, myocardial infarction, congestive heart failure, or stroke.^16^ An observational study using the target trial approach in 22,474 dialysis patients aged ≥65 years with diabetes and cardiovascular disease from the United States Renal Data System (USRDS) evaluated clinical strategies for managing anaemia with epoetin therapy. ^26^ The study found similar 6-month survival targeting a medium haematocrit range (34.5%–39.0%) and a lower haematocrit range (30.0%–34.5%), HR 0.98 (95% CI 0.78– 1.24). A further analysis of USRDS data using the target trial approach with the parametric g-formula found that, compared to a low-haematocrit strategy (30–33%), the estimated risk of death was 4.6% higher (95% CI: 4.4–4.9) under a high-haematocrit strategy (36–39%) and 1.8% higher (95% CI: 1.7–1.9) under a mid-haematocrit strategy (33–36%).^27^ This study used an 18-month follow-up and included patients with a history of congestive heart failure or ischaemic heart disease in the two years prior to study entry.

Previous analyses of UKRR data have shown that anaemia management patterns for haemodialysis patients in the UK changed considerably between 2005 and 2013.^28^ There was a decrease in ESA use, the average dose administered, and the achieved Hb levels in UK haemodialysis patients over time. These trends suggest that the results from randomized trials like CHOIR, CREATE, and TREAT, along with updated clinical recommendations, have influenced clinical decision-making, prompting a more conservative approach to anaemia management in haemodialysis patients. The Proactive IV Iron Therapy in Haemodialysis Patients (PIVOTAL) trial, published after our data extraction period, is likely to have further influenced clinical practice by demonstrating that a proactive high-dose intravenous iron regimen reduced ESA requirements and improved clinical outcomes compared to a reactive low-dose approach.^29^ As a result, current ESA prescribing is often combined with diverse iron strategies, which can complicate the interpretation of ESA effects alone. By analysing data that precede the PIVOTAL trial, our study offers a more isolated assessment of ESA treatment strategies, minimising influence from evolving iron management practices. Our results suggest that a modestly higher Hb target may not be detrimental to patient survival, providing that excessive ESA dosing is avoided.

A strength of our analysis was the availability of detailed observational data, which allowed us to emulate a target trial and assess dynamic treatment strategies reflecting clinical practice. The UKRR is a large and representative database allowing trends in clinical practice patterns to be captured.^30^ By structuring the data into monthly time periods we were able to incorporate information on Hb levels that informed darbepoetin treatment decisions. The study design utilised variability in practitioners’ usual darbepoetin dosing practices over time, assuming this variation was not systematically linked to other, unmeasured, factors influencing mortality. However, our study was limited by incomplete data returns, as some kidney centres do not routinely record computerised data on ESA dose or drug type. We therefore restricted analyses to centres where at least 60% of haemodialysis patients were reported as receiving ESA treatment. A key limitation of this study is its observational nature: we cannot exclude unmeasured confounding. However, we controlled for the key variables that predict treatment decisions and/or are associated with all-cause mortality.

In conclusion, our target trial emulation using EHR data found no evidence that targeting a Hb range of 105– 125 g/L, compared with a 95–115 g/L range, with a maximum darbepoetin dose of 150 µg/week, increases all-cause mortality at 8 months in haemodialysis patients. However, targeting a range of 125–145 g/L may increase mortality compared with a 95–115 g/L range. Haemodialysis patients may benefit from modestly higher Hb, in terms of symptoms and quality of life, under a dosing strategy that limits changes in dose and maximum permissible dose of ESAs.

## Disclosures / Conflict of interest statement

None to declare.

## Data sharing statement

The dataset is not available due to privacy restrictions. Researchers may apply for access to UK Renal Registry data through the UK Kidney Association. Information and application procedures is provided at https://www.ukkidney.org/audit-research/how-access-data.

## Acknowledgements

We thank all the UK kidney centres for providing data to the UK Renal Registry. We thank Miguel Hernán for comments on the analyses. We thank Charles Thompson for his input into the original study idea and the development of the trial design.

## Funding

KB was supported by a Medical Research Council (MRC) UK fellowship (R137881-101). YBS is partly funded by National Institute for Health and Care Research Applied Research Collaboration West (NIHR ARC West) and University of Bristol.

## Author contributions

JACS, KB, FJC, YBS and DN designed the study. FJC and AC advised on the data. KB performed the analyses. JACS and EJM advised on the analyses. All authors contributed to interpreting results. KB wrote the initial paper draft and all other authors edited and provided feedback on drafts. All authors read and approved the final manuscript.

## Notes

### Competing Interest Statement

The authors have declared no competing interest.

