## Supplementary Information for "Evaluating treatment strategies for managing anaemia with erythropoiesis stimulating agent therapy in haemodialysis patients: findings from a target trial emulation using electronic health record data"

**Figure A1 Acceptable dose changes (μ/week)**

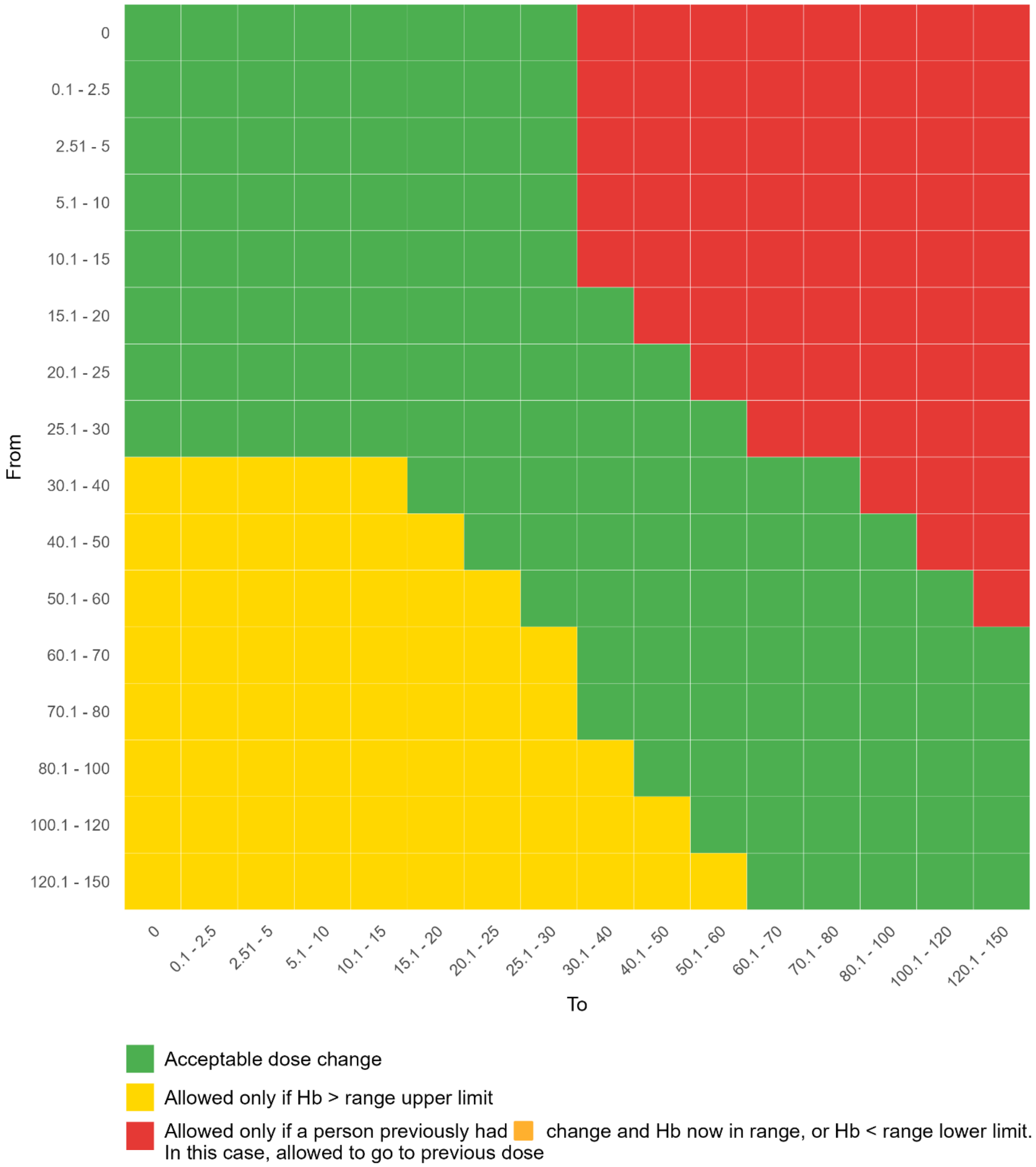

### Figure A2 Flow chart of patients included in the study

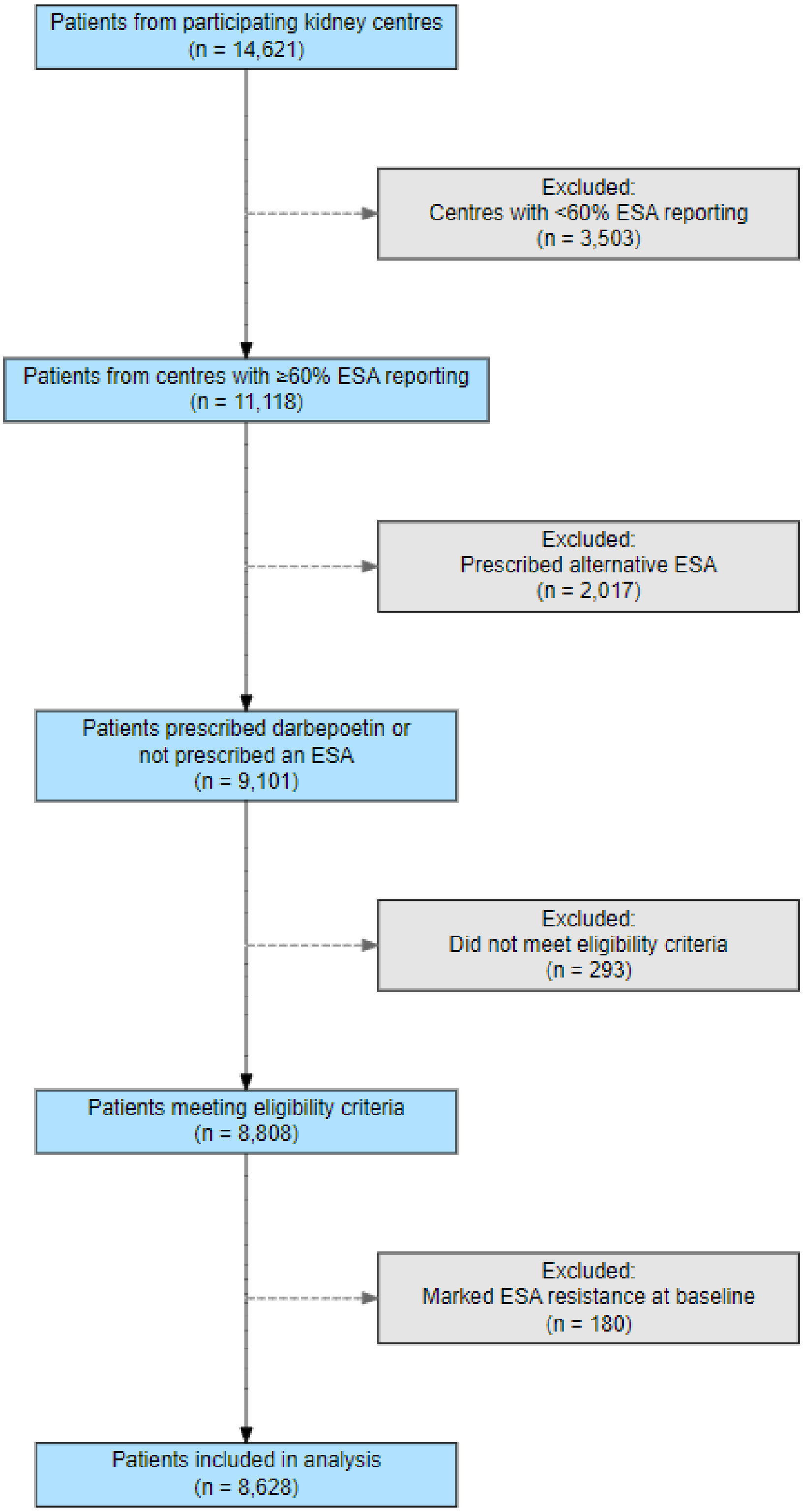


Notes
ESA: Erythropoiesis-Stimulating Agent
